# Targeted Immunosuppression Distinguishes COVID-19 from Influenza in Moderate and Severe Disease

**DOI:** 10.1101/2020.05.28.20115667

**Authors:** Philip A. Mudd, Jeremy Chase Crawford, Jackson S. Turner, Aisha Souquette, Daniel Reynolds, Diane Bender, James P. Bosanquet, Nitin J. Anand, David A. Striker, R. Scott Martin, Adrianus C. M. Boon, Stacey L. House, Kenneth E. Remy, Richard S. Hotchkiss, Rachel M. Presti, Jane A. O’Halloran, William G. Powderly, Paul G. Thomas, Ali H. Ellebedy

## Abstract

Coronavirus disease 2019 (COVID-19) is characterized by a high incidence of acute respiratory failure. The underlying immunopathology of that failure and how it compares to other causes of severe respiratory distress, such as influenza virus infection, are not fully understood. Here we addressed this by developing a prospective observational cohort of COVID-19 and influenza subjects with varying degrees of disease severity and assessing the quality and magnitude of their immune responses at the cellular and protein level. Additionally, we performed single-cell RNA transcriptional profiling of peripheral blood mononuclear cells from select subjects. The cohort consists of 79 COVID-19 subjects, 26 influenza subjects, and 15 control subjects, including 35 COVID-19 and 7 influenza subjects with acute respiratory failure. While COVID-19 subjects exhibited largely equivalent or greater activated lymphocyte counts compared to influenza subjects, they had fewer monocytes and lower surface HLA-class II expression on monocytes compared to influenza subjects and controls. At least two distinct immune profiles were observed by cytokine levels in severe COVID-19 patients: 3 of 71 patients were characterized by extreme inflammation, with greater than or equal to ∼50% of the 35 cytokines measured greater than 2 standard deviations from the mean level of other severe patients (both influenza and COVID-19); the other immune profile, which characterized 68 of 71 subjects, had a mixed inflammatory signature, where 28 of 35 cytokines in COVID-19 patients had lower mean cytokine levels, though not all were statistically significant. Only 2 cytokines were higher in COVID-19 subjects compared to influenza subjects (IL-6 and IL-8). Influenza and COVID-19 patients could be distinguished statistically based on cytokine module expression, particularly after controlling for the significant effects of age on cytokine expression, but again with lower levels of most cytokines in COVID-19 subjects. Further, high circulating levels of IL-1RA and IL-6 were associated with increased odds of intubation in the combined influenza and COVID-19 cohort [OR = 3.93 and 4.30, respectively] as well as among only COVID-19 patients. Single cell transcriptional profiling of COVID-19 and influenza subjects with respiratory failure identified profound suppression in type I and type II interferon signaling in COVID-19 patients across multiple clusters. In contrast, COVID-19 cell clusters were enriched for alterations in metabolic, stress, and apoptotic pathways. These alterations were consistent with an increased glucocorticoid response in COVID-19 patients compared to influenza. When considered across the spectrum of innate and adaptive immune profiles, the immune pathologies underlying severe influenza and COVID-19 are substantially distinct. The majority of COVID-19 patients with acute respiratory failure do not have a cytokine storm phenotype but instead exhibit profound type I and type II IFN immunosuppression when compared to patients with acute influenza. Upregulation of a small number of inflammatory mediators, including IL-6, predicts acute respiratory failure in both COVID-19 and influenza patients.

## Introduction

Infection with severe acute respiratory syndrome coronavirus 2 (SARS-CoV-2) causes coronavirus disease 2019 (COVID-19). Acute respiratory failure occurs in a subset of COVID-19 patients.(*1–3*) Respiratory failure has occurred in as many as 8% of individuals testing positive for infection in the Lombardy region of Italy.(*4*) Understanding the etiology of respiratory failure in COVID-19 patients is critical for determining the best management strategies and pharmacologic targets for treatment. Current management of acute respiratory failure in COVID-19 consists of optimized supportive care,(*5, 6*) primarily through oxygen administration and consideration of endotracheal intubation and mechanical ventilation in the appropriate context.(*7*)

Cytokine storm syndrome (CSS) is increasingly proposed as underlying the etiology of respiratory failure in patients with COVID-19.(*8*) This model suggests that respiratory failure is related to significant pro-inflammatory cytokine expression that leads to inflammatory cell recruitment and tissue damage in the lung. Most of the data supporting this hypothesis in COVID-19 comes from an early paper that observed high levels of the cytokines IL-2, IL-7, IL-10, GCSF, IP-10, MCP-1, MIP-1α and TNFα in a small cohort of COVID-19 patients cared for in the ICU. The level of these cytokines was increased in the ICU patients compared with a group of COVID-19 patients that did not require care in the ICU.(*3*)

There has been significant interest in modulating the systemic immune response in an effort to prevent or treat respiratory failure in patients with COVID-19.(*2, 9–11*) More than one hundred clinical trials are currently registered at clinicaltrials.gov to evaluate the efficacy of inflammatory cytokine blocking medications or interventions such as cytokine filtration as potential treatments for respiratory failure in COVID-19 patients. A thorough understanding of the underlying inflammatory environment in COVID-19 patients is required to successfully interpret the findings of these studies.

Acute respiratory failure in influenza patients has also been attributed to significantly increased inflammation and CSS.(*12–14*) In order to evaluate the etiology of respiratory failure in COVID-19 patients, we undertook a comparative investigation of inflammatory responses in a cohort of influenza patients with severe illness collected during 2019 and 2020, which allowed us to characterize the immune response in patients with severe COVID-19 specifically in the context of the more widely studied immune responses seen during influenza disease.

## Results

### Demographic and Clinical Characteristics

We enrolled a total of 79 symptomatic subjects who tested positive for SARS-CoV-2 RNA using an FDA-approved clinical PCR test. Our comparison cohort consisted of 26 symptomatic influenza subjects recruited during the 15 months immediately preceding the outbreak of COVID-19 in the Saint Louis region. All tested positive for influenza A or B via a clinical PCR test obtained during their clinical care. COVID-19 subjects were significantly older than both influenza and control subjects, and significantly more COVID-19 patients required hospitalization (Table 1). A higher number of COVID-19 subjects required ICU admission and mechanical ventilation than influenza subjects, but this was not significantly different. Twenty-seven percent of the COVID-19 subjects died during their hospitalization, compared with 8% of influenza subjects enrolled. Less than 5% of the combined COVID-19 and influenza cohort were immunocompromised. Many subjects in both cohorts exhibited co-morbidities that increased their risk for severe disease, including diabetes and chronic lung disease; however, there were no significant differences between the COVID-19 and influenza subjects in any measured comorbidity (Table 1).

**Table 1.**
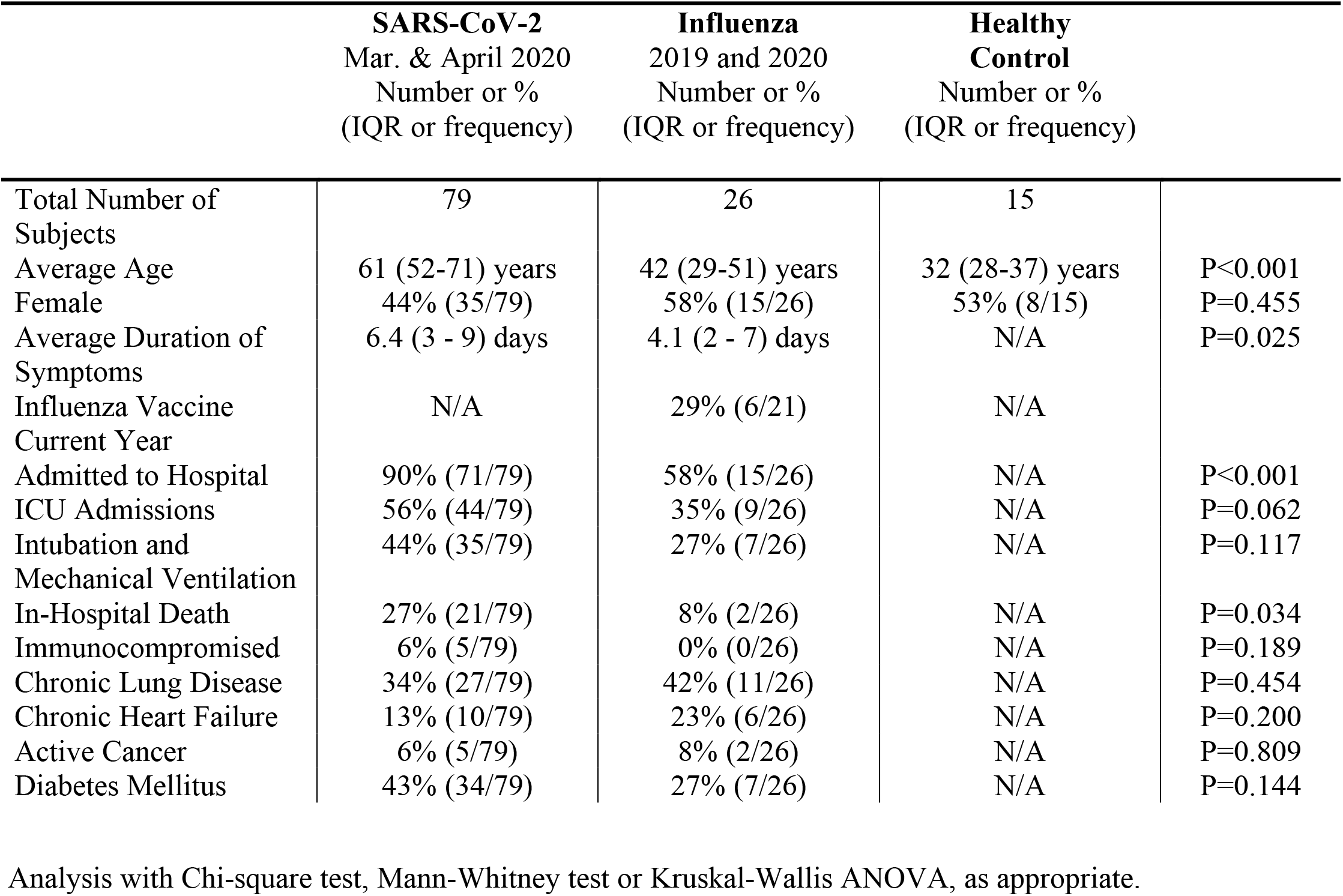
Clinical cohort characteristics.

|  | **SARS-CoV-2**<br>Mar. & April 2020<br>Number or %<br>(IQR or frequency) | **Influenza**<br>2019 and 2020<br>Number or %<br>(IQR or frequency) | **Healthy Control**<br>Number or %<br>(IQR or frequency) |  |
| --- | --- | --- | --- | --- |
| Total Number of Subjects | 79 | 26 | 15 |  |
| Average Age | 61 (52-71) years | 42 (29-51) years | 32 (28-37) years | P<0.001 |
| Female | 44% (35/79) | 58% (15/26) | 53% (8/15) | P=0.455 |
| Average Duration of Symptoms | 6.4 (3 - 9) days | 4.1 (2 - 7) days | N/A | P=0.025 |
| Influenza Vaccine Current Year | N/A | 29% (6/21) | N/A |  |
| Admitted to Hospital | 90% (71/79) | 58% (15/26) | N/A | P<0.001 |
| ICU Admissions | 56% (44/79) | 35% (9/26) | N/A | P=0.062 |
| Intubation and Mechanical Ventilation | 44% (35/79) | 27% (7/26) | N/A | P=0.117 |
| In-Hospital Death | 27% (21/79) | 8% (2/26) | N/A | P=0.034 |
| Immunocompromised | 6% (5/79) | 0% (0/26) | N/A | P=0.189 |
| Chronic Lung Disease | 34% (27/79) | 42% (11/26) | N/A | P=0.454 |
| Chronic Heart Failure | 13% (10/79) | 23% (6/26) | N/A | P=0.200 |
| Active Cancer | 6% (5/79) | 8% (2/26) | N/A | P=0.809 |
| Diabetes Mellitus | 43% (34/79) | 27% (7/26) | N/A | P=0.144 |

Analysis with Chi-square test, Mann-Whitney test or Kruskal-Wallis ANOVA, as appropriate.

### Evaluation of Circulating Immune Cells

We initially characterized circulating immune cells by quantifying the absolute number of CD4^+^ and CD8^+^ T lymphocytes and CD19^+^ B cells. COVID-19 and influenza subjects both exhibited universally reduced populations of these three cell subsets, which generally constitute the majority of circulating PBMCs in healthy controls (Fig 1A, Supplemental Fig 1A). In contrast, COVID-19 subjects had significantly more circulating early antibody-secreting B cell plasmablasts than influenza subjects or controls (Fig 1B). Circulating activated CD8 T cells were equivalent across all groups, but circulating activated CD4 T cells were significantly lower in influenza compared healthy controls (Fig 1B). However, when compared with either influenza or control subjects, COVID-19 subjects exhibited significantly reduced numbers of circulating monocytes, including all three common classifications of human monocytes (classical, intermediate, and non-classical; Fig 1C, Supplemental Fig 1B). In line with previous observations(*15*), intermediate monocytes were elevated during acute influenza in comparison to healthy controls.

**Figure 1.**
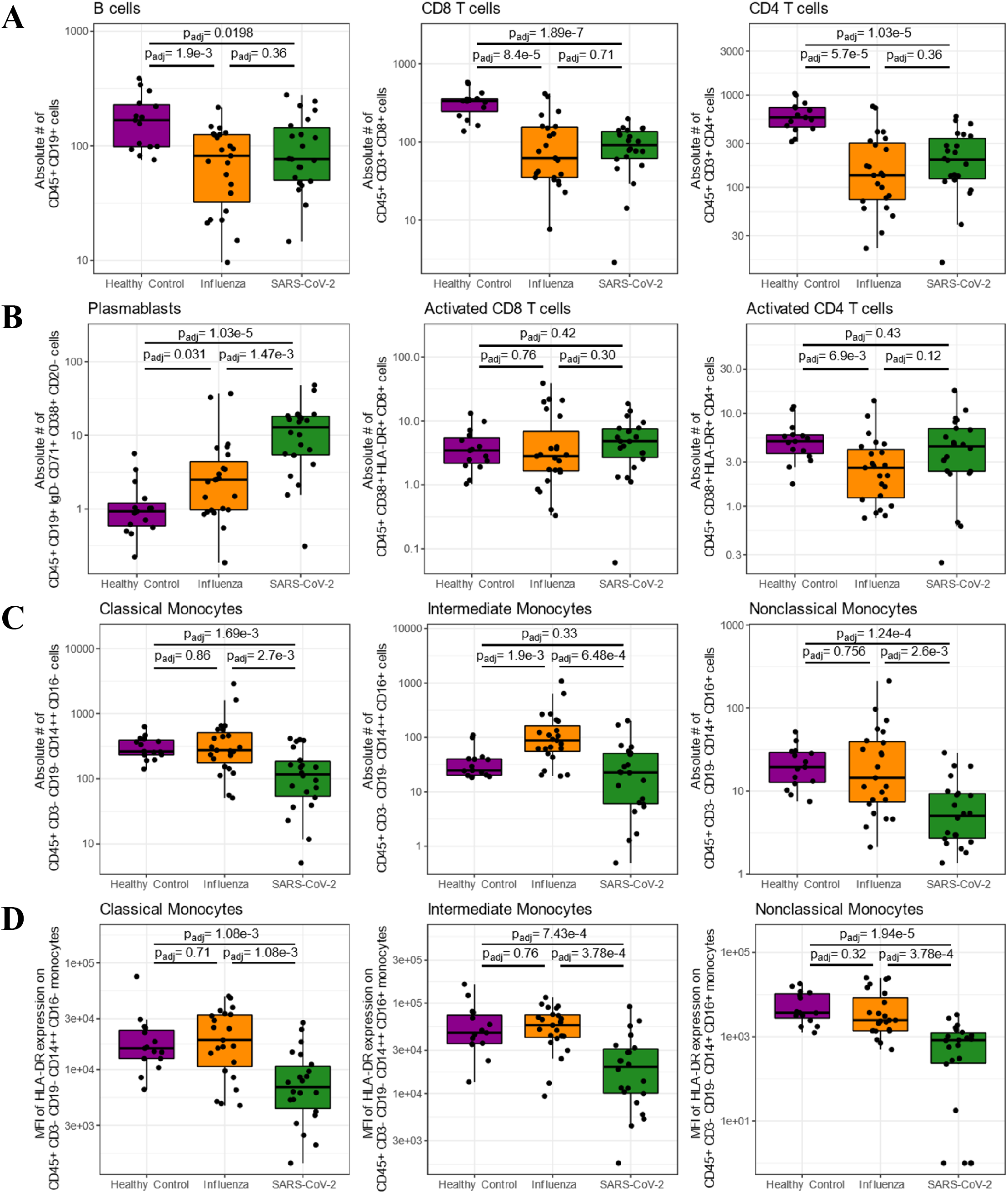
Evaluation of absolute numbers of circulating lymphocyte and monocyte subpopulations in select healthy controls (N = 15), acute influenza-infected subjects (N = 23), and acute SARS-CoV-2-infected subjects (N = 22). **(A)** Total B cells, CD8 and CD4 T cells; **(B)** circulating B cell plasmablasts, activated CD8 and CD4 T cells; and **(C)** classical, intermediate, and nonclassical monocytes. **(D)** Surface expression of the major histocompatibility complex class 2 molecule, HLA-DR, on the surface of the indicated sub-populations of circulating monocytes as measured by geometric mean florescent intensity (MFI) using flow cytometry. Presented p-values are from Mann-Whitney U tests, post adjustment for multiple testing.

Given the pronounced variation in monocyte abundance across patient conditions, we also measured major histocompatibility complex class II expression on the surface of monocytes to gauge monocyte activation. We noted that COVID-19 subjects had significantly reduced abundances of HLA-DR on the surface of all three subsets of monocytes when compared with either controls or influenza subjects (Fig 1D). There was no significant difference in the cell-surface expression of HLA-DR on B cells or CD8 T cells, but COVID-19 subjects exhibited significantly less HLA-DR expression on CD4 T cells compared to influenza subjects (Supplemental Fig 1C).

### Cytokine associations with disease and outcome

Plasma cytokine levels were measured from 27 patients with confirmed influenza virus infection, 79 patients with SARS-CoV-2 infection, and 8 healthy controls. Among the SARSCoV-2 patients, two response profiles were immediately apparent. Three of 79 patient samples had extremely high concentrations, defined as > 2 standard deviations from the mean, for more than 17 of the 35 cytokines measured (range: 49%-89%), characteristic of a classic cytokine storm (see outliers, Supplemental Fig 2). Standard deviations from the mean ranged from 2 up to 10.5 among these subjects, with outlier values ranging from 0.8 to 2 orders of magnitude higher than the mean for each of the measured cytokines. We excluded these 3 patients from subsequent comparative analyses.

The dominant response profile among COVID-19 patients consisted of more selective cytokine upregulation, with a bias towards lower inflammation when compared to influenza patients (Fig 2A). Among all subjects, we found that for 28 of 35 cytokines, COVID-19 patients had lower mean cytokine levels, though not all were statistically significant (Supplemental Fig 3). Among the statistically significant reduced cytokines were GM-CSF (padj = 0.0499), IFN-γ (padj = 0.038), and IL-9 (padj = 0.026, Figure 2B). In contrast, only IL-6 (padj = 0.086) and IL-8 (padj = 0.01) were near significantly or significantly elevated in the total COVID-19 group compared to all influenza patients (Figure 2B). Cytokines upregulated similarly in COVID-19 and influenza during infection included chemokines (IP10, MIP1β, MCP1, MIG) and immunomodulatory cytokines (HGF and IL1-Ra, Supplemental Fig 3). These data indicated that the majority of COVID-19 patients did not have as profound an inflammatory phenotype as influenza patients, with certain targeted exceptions.

**Figure 2.**
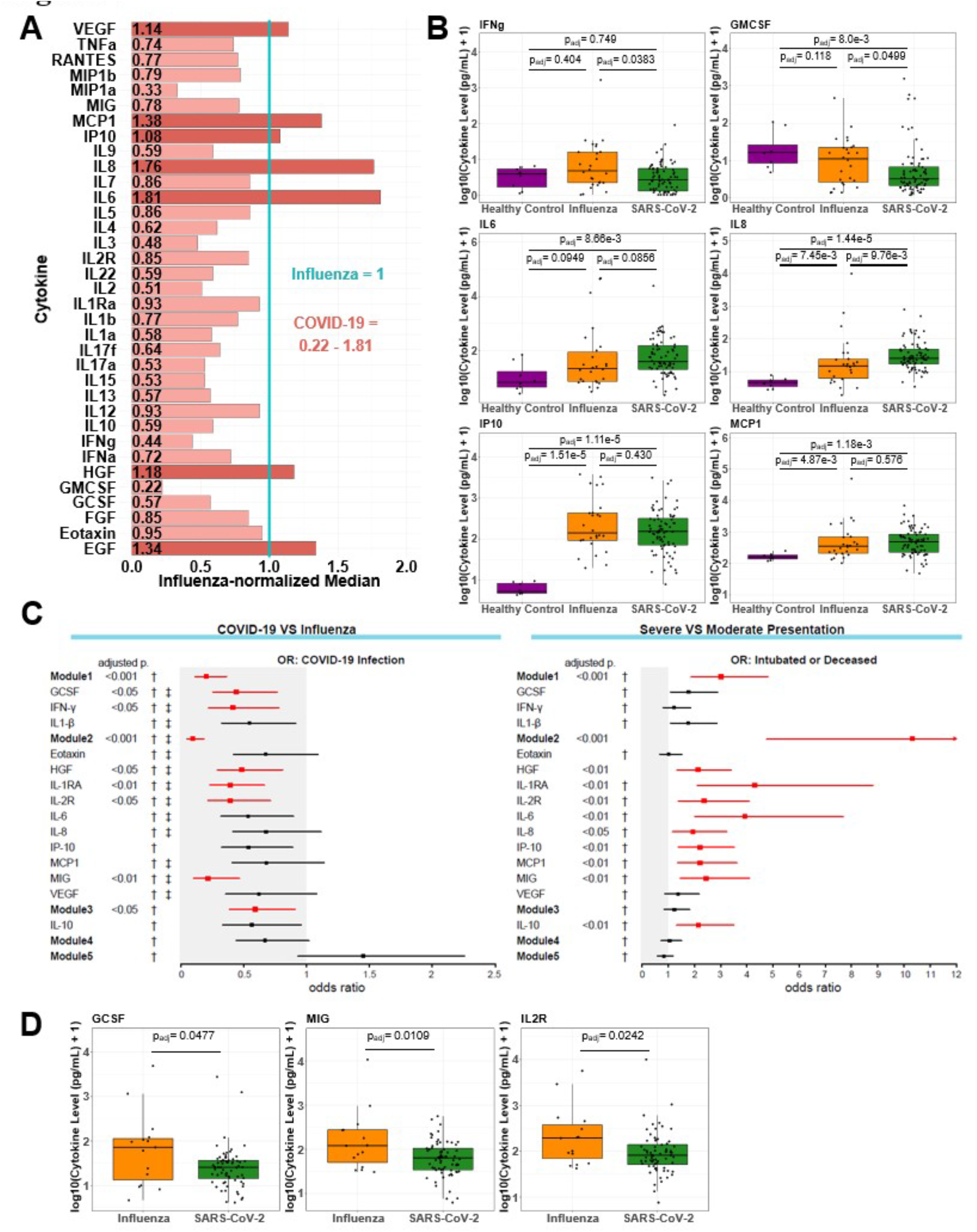
Selective cytokine upregulation in COVID-19 patients. **(A)** To visualize cytokine levels in COVID-19 patients relative to influenza patients, all cytokine levels were normalized to the median cytokine level in influenza subjects, with respect to cytokine; thus, the normalized median cytokine level in influenza patients equals 1 for all cytokines and is represented by the vertical blue line. Bar graphs, and the numbers on them, represent the value of the normalized median COVID-19 cytokine level relative to the normalized median influenza cytokine level for a given cytokine. Bars in light red represent cytokine levels lower in COVID-19 patients relative to influenza patients (normalized median < 1, n = 28) and bars in dark red are cytokines higher in COVID-19 patients relative to influenza patients (normalized median > 1, n = 7). **(B)** Box plots show cytokine concentrations in COVID-19, influenza, and healthy subjects: Presented p-values are from post hoc Dunn’s tests, following adjustment for multiple testing. **(C)** Forest plots depicting the odds ratios obtained from logistic regression analysis between cytokines and COVID-19 infection (left) and severe disease (i.e., resulting in intubation or death; right). Logistic regression models utilized absolute log_10_-transformed cytokine values and included age and the number of days since symptom onset at sampling as covariates. Results from each cytokine module are presented, as well as results from each constituent cytokine Modules 1 and 2, which showed the most variation across tested outcomes; otherwise, cytokines are only presented if they reached statistical significance in one of the logistic regression models. Grey shading indicates the area of the plots where odds ratios are less than 1, which are indicative of negative associations. Presented p-values correspond to the specific module or cytokine and were adjusted for multiple testing. Odds ratios are indicated with points, and confidence lines encompass the range between the lower and upper limits. Red points and confidence lines indicate FDR-adjusted p-values < 0.05. For each model, † indicates that age was a significant covariate, whereas ‡ indicates that day of sampling was statistically significant. **(D)** Cytokine concentrations in hospitalized COVID-19 and influenza subjects. Presented p-values are based on logistic regression analysis in the CytoMod package, following adjustment for multiple testing.

We and others have previously shown that cytokine levels are often correlated within a subject based on demographic and environmental factors (e.g., age, prior herpesvirus exposure). This can obscure variation in cytokine expression patterns within individuals. Further, the relatively high dimensionality of cytokine data can make it difficult to detect statistically significant associations due to high false discovery rates. Additionally, the distinct distributions in our cohorts between severe and mild patients complicate simple overall comparisons. To address this, we utilized a data-driven modular informatics approach(*17*) to identify clusters of co-regulated cytokines across all subjects, to normalize cytokine expression within subjects, and to identify differences in expression across conditions while adjusting for the effects of age and days after symptom onset for sampling. These analyses allowed us to detect a number of co-correlating cytokines across COVID-19 and influenza samples, which we grouped into distinct co-expression modules using hierarchical clustering (Supplementary Figure 4). Of particular interest were Modules 1 and 2, which included GCSF, IFN-γ, and IL1-β (MOD1) and Eotaxin, HGF, IL1-Ra, IL-2R, IL-6, IL-8, IP10, MCP1, MIG, and VEGF (MOD2), respectively. Upon constructing logistic models to assess associations between these cytokines and a binary infection status (COVID-19 vs. influenza), we observed that *lower* levels of both MOD1 and MOD2, as well as a number of their constituent cytokines, were significantly associated with an *increased* risk of being COVID-19 positive (Fig 2A). This was particularly surprising given that IL-6 and IL-8 are frequently associated with severe COVID-19 infection and were upregulated in COVID-19 compared to influenza subjects in the present study.

However, the modular analysis considers all of the cytokines within a module, as opposed to individual cytokine associations, and MOD2 contains several other cytokines; indeed, neither IL-6 nor IL-8 alone were significantly associated with being COVID-19 positive. Furthermore, the logistic model specifically regresses out potentially confounding covariates that are otherwise obscured by simple univariate analysis. These results suggest that overall higher inflammation is foremost predictive of influenza infection and that a defining feature of COVID-19 disease is generally reduced inflammation compared to influenza as measured by a number of related cytokines. Thus, influenza infection was associated with generally higher levels of inflammation, and when influenza subjects exhibited high expression in one MOD2 cytokine, they were likely to exhibit high expression across the board. In contrast, COVID-19 subjects were not characterized by overall high levels of cytokines, but rather exhibited a selective pattern of inflammation, in which only a subset of inflammatory cytokines are upregulated.

Using this analytical framework further, we next inquired whether particular cytokine expression patterns were associated with poor clinical outcomes. In considering all infected subjects, higher levels of Modules 1 and 2 were significantly associated with severe disease, which we defined as resulting in either intubation or death. Associations between disease severity and IL1-RA and IL-6 were particularly strong, with high levels of these analytes predictive of poor outcomes (OR = 3.93 and 4.30, respectively; Fig 2C). Importantly, this association remained when we repeated the analysis on COVID-19 subjects alone (Supplementary Figure 5), demonstrating that exacerbation of these specific inflammatory pathways in COVID-19 in the sickest patients.

Because these earlier analyses included severity as an outcome within a population of patients with diverse disease presentations, we also tested the robustness of our observations specifically in hospitalized patients from both groups. Once again, the striking pattern that emerged was lower levels of multiple cytokines and modules among hospitalized COVID-19 patients compared to hospitalized influenza patients. These included GCSF (padj = 0.048), MIG (padj = 0.011), and IL-2R (padj = 0.024), and spanned three distinct cytokine modules. Taken together, COVID-19 is characterized by high levels of specific cytokines (IL-6, IL-8) when compared to influenza, but the majority of inflammatory mediators are significantly lower, even among severe patients (Fig 2C-D), especially after accounting for the frequently significant effect of patient age on cytokine abundance.

### Single Cell Transcriptional Profiles of Subjects with Respiratory Failure Reveals Immunosuppression within Monocytes and Increased Glucocorticoid Response in COVID-19 Subjects

Immune suppression can often occur as a negative feedback from immune activation, so we sought further resolution of the immune state of severe COVID-19 patients to understand the dominant regulatory signals determining their trajectory. A total of 29,634 cells from seven subjects (three COVID-19, three influenza, and one healthy control) were obtained for single cell gene expression analyses after standard processing and filtering. Using an integration-based approach that leverages convergent expression signals across samples (see Methods), we identified 22 putative transcriptional clusters that we were able to categorize into major cell subsets, including monocytes and macrophages (7 transcriptional clusters, including CD16+ and CD16-monocytes), CD8+ T cells (5 clusters), CD4+ T cells (3 clusters), B cells (2 clusters), regulatory T cells (Tregs), mixed cytolytic lymphocyte populations (MCLPs; e.g., NK and NKT cells), plasmablasts, mast cells, and three other subsets that we putatively identified as plasmacytoid dendritic cells (PDCs), platelets, and cell doublets composed in part of platelets. We then interrogated each of these major groups and their constituent transcriptional clusters for variation in both relative abundance and gene expression owed to differences in condition (i.e., COVID-19-infected, influenza-infected, or healthy control). Although representation of cells within the clusters were influenced by their abundance in individual patients (Supplementary Figure 6), the models we leveraged to compare across conditions are robust to such differences, emphasizing transcriptional variation between conditions among similar subsets.

To broadly survey transcriptional variation as a function of infection status in an unbiased manner, we ranked gene expression differences between COVID-19-infected and influenza-infected patients for each subset and tested for enrichment of Hallmark gene sets as a function of these diagnoses. Surprisingly, a number of important immunological pathways were significantly enriched specifically among cells from Influenza patients across a number of subsets: compared to the Influenza condition, both IFN-γ and IFN-α response pathways were significantly downregulated within the COVID-19 condition for B cells, CD8+ T cells, MCLPs, Tregs, PDCs, and monocyte/macrophage subsets (Fig 3E; Supplementary Figure 7). More exhaustive analysis using gene ontology pathways related to interferon production, secretion, response, and regulation demonstrated that these patterns extended to IFN-β and general Type I interferon pathways across most subsets, but particularly among monocytes (Supplementary Figure 8). These patterns were concordant with substantial enrichment of inflammatory pathways in Influenza cells compared to COVID-19 cells across a majority of cell subsets. In contrast, COVID-19 cells were significantly enriched for a number of pathways involved in cellular metabolism and proliferation in comparison to Influenza cells across most subsets (Supplementary Figure 7).

**Figure 3.**
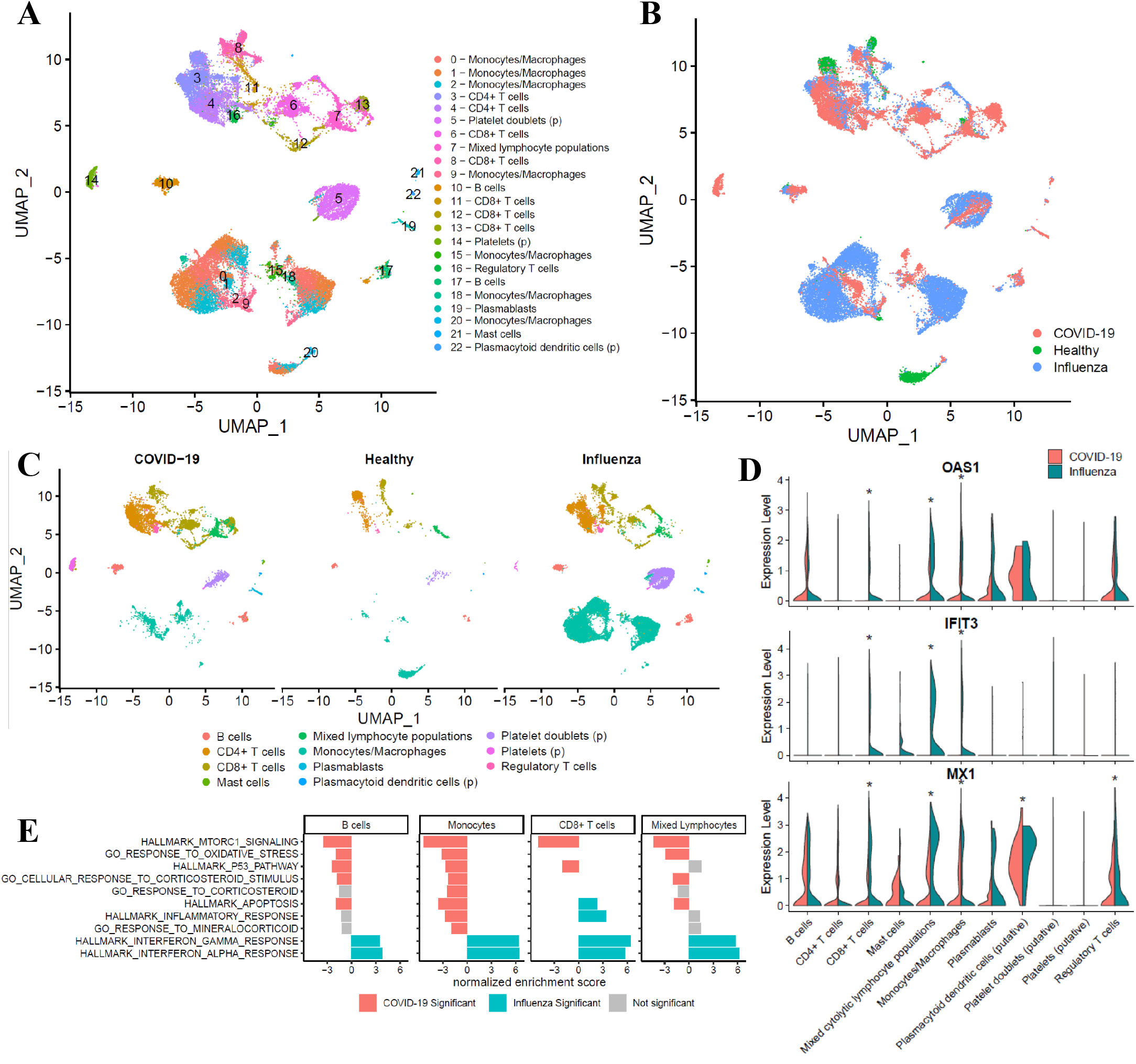
Single-cell gene expression analyses of PBMCs from COVID-19-positive, influenza-positive, and healthy subjects demonstrate profound differences in the relative abundance and transcriptional activity of cell subsets across conditions. **(A)** UMAP (Uniform Manifold Approximation and Projection) plots depict transcriptional clusters, which **(B)** vary transcriptionally as a function of condition despite the presence of nearly all subsets across the various conditions, as evidenced in **(C)**. Putative cell subset identities are denoted with (p). **(D)** Violin plots demonstrate extreme downregulation of selected interferon-activated (OAS1) or interferon-induced (IFIT3, MX1) genes among cells from COVID-19 patients (red) compared to cells from influenza patients (blue). Asterisks indicate significance at FDR < 0.05. **(E)** In direct comparison to cells from influenza-infected patients, transcriptional patterns among cells from COVID-19 patients reveal significant upregulation (red bars) of metabolic pathways, stress pathways, and glucocorticoid signaling pathways, particularly in monocytes/macrophages; in contrast, interferon pathways were significantly downregulated (blue bars) among subsets from COVID-19 patients. Grey bars indicate that tests for enrichment did not meet statistical significance for a particular subset.

Given the substantial downregulation of HLA-DR among COVID-19 monocytes observed during flow cytometry analysis, we decided to further investigate potential transcriptional differences between COVID-19 and Influenza specifically within our transcriptionally defined monocyte/macrophage subset and clusters. As expected given the flow cytometry analysis carried out on these same patients, the proportion of cells in monocyte/macrophage subsets were substantially smaller in COVID-19 compared to both healthy and influenza subjects (Supplementary Figure 9). We furthermore noted significant decreases in a number of interferon-induced genes known to play important roles in the innate immune response to viral insults (Fig 3D). This evidence of decreased, or perhaps dysregulated, interferon expression and a relative dearth of expression corresponding to inflammatory processes were accompanied by a significant enrichment of a number of stress and corticoid response pathways in COVID-19 cells across most subsets, but particularly within monocytes/macrophages (Fig 3E; Supplementary Figure 10).

## Discussion

Understanding the complexities of the systemic inflammatory response to SARS-CoV-2 infection is critical to determining the most appropriate treatment for this condition. We have demonstrated that the immunophenotype of COVID-19 and influenza patients vary widely. Two forms of COVID-19 immune dysregulation were observed: a cytokine storm phenotype in a small proportion of patients (3 of 79) and a far more common phenotype characterized by targeted immunosuppression. The signatures of this common COVID-19 phenotype were highly elevated IL-6 and IL-8, paired with lower levels (compared to severe influenza) of cytokines in many other pathways and essentially the absence of any Type I or Type II IFN response. The suppression in Type I IFN signaling has been noted by others in humans and animal models of COVID-19 infections(*18, 19*). At the cellular level, dramatic reductions in overall cellularity and particularly in the monocyte compartment were observed, with phenotypic and transcriptional evidence (Class II downregulation) that monocytes were less activated. While lymphocyte numbers (except for plasmablasts) were reduced in both infected groups compared to healthy controls, several lymphocyte subsets had functional signatures of suppression in COVID-19 patients, including type I and II IFN signaling. IFN-γ production is critical for effector type I responses, and its absence may limit antiviral activity. The elevated plasmablast frequencies in COVID-19 patients may reflect the abundance of viral antigens, which is consistent with the reported persistence of viral RNA in nasal swabs for up to 15 days after onset of symptoms(*20*).

The single cell analyses also identified enrichment of several pathways in COVID-19 patients associated with metabolic stress and the general stress response. These results, combined with the targeted, severe suppression of specific pathways and dramatic leuko- and lymphopenia led us to consider what pathways might account for this response profile. Previous studies in animal models had implicated glucocorticoid (GC) signaling in the immunosuppression and lymphopenia that occurs in the influenza model in mice.(*21*) In humans, systemic inflammation, and hydrocortisone specifically, are known to suppress HLA-DR expression on monocytes.(*22*)

Excessive GC production is an attractive hypothesis to account for the observed immune dysregulation and disease manifestations in COVID-19. First, the high levels of IL-6 production—one of the two cytokines that were higher in COVID-19 than in severe influenza— can directly drive excessive cortisol production through multiple mechanisms, including through the direct induction of corticotropin-releasing hormone and adrenocorticotropin. IL-6 can also act directly on the adrenal cortex to stimulate GC release.(*23*) While GCs are generally immunosuppressive, which is why they are often considered therapeutically, their effects are uneven across the cytokine landscape, with cortisol failing to suppress IL-6(*24*), or even inducing IL-6 and IL-8 in one report.(*25*) Notably, in this study there were differences in cortisol’s effects between male and female cells, with more pro-inflammatory effects observed in males; male sex has been noted as a consistent risk factor in COVID-19.(*26*) A recent retrospective cohort study from Germany has also reported increased cortisol levels in a majority of COVID-19 patients.(*27*)

Another potential source of cortisol production unique to SARS-CoV-2 infection is through its modulation of ACE2 levels. ACE2 is the receptor for virus entry, and while it is still unclear the extent to which its levels are modulated by infection, experience from SARS-CoV-1 and other data suggest that ACE2 expression or activity are lowered. This directly affects the renin-angiotensin system, enhancing ACE activity and resulting in a shift towards angiotensin II (Ang II) production and away from angiotensin 1–7. Ang II enhances Il-6 and cortisol production, while Ang 1–7 suppresses them.(*28, 29*)

Beyond considering the potential impacts of increased GC production on the development of the immunosuppressed phenotype we characterized in the vast majority of COVID-19 patients in this study, we must also consider the possibility that GCs modulate the immune microenvironment differently in the context of distinct viral encounters. There are known but underappreciated immunosuppressive features of influenza (*30–32*); coronaviruses are similarly known to utilize a number of nonstructural proteins to disrupt host gene expression and protein synthesis (*33, 34*), with the potential for profound impacts on various immune signaling pathways (*35*). Although the details of these mechanisms in the context of COVID-19 immune dysregulation remain to be fully elucidated, our own analyses demonstrate enhanced GC signaling in cells from COVID-19 patients, consistent with an enhanced production of GCs or enhanced sensitivity to GCs after infection. Together these data suggest a feedforward amplification of IL-6 and GC signaling, paired with a profound suppression of other potentially protective immune functions through GC-induced apoptosis and suppression of key antiviral pathways. GCs can also drive other pathological phenotypes including clotting dysfunction, consistent with severe manifestations of COVID-19.(*36*) The direct and indirect effects of SARS-CoV-2’s unique biology may determine this pathological outcome. Therapeutic consideration should be given to inhibiting both IL-6 and GC activity in the majority of COVID-19 patients exhibiting this phenotype (high IL-6, low IFN signaling, profound cytopenias) versus the small proportion of patients with a true cytokine storm phenotype. While inhibiting GC activity alone may cause excessive inflammation, co-administration with IL-6-blocking antibodies in a dose regulated manner may limit these effects. These data also indicate that administration of GCs would potentially be deleterious, an outcome consistent with some limited reports.(*37*) Other studies have indicated beneficial or neutral effects from GC, and indicate a focus on understanding their dynamics and activity across the course of infection is an important direction for future research.(*38, 39*)

## Materials and Methods

### Study Design

This is a prospective observational cohort of subjects with viral respiratory illness symptoms who presented to Barnes Jewish Hospital, St. Louis Children’s Hospital, Missouri Baptist Medical Center or affiliated Barnes Jewish Hospital testing sites located in Saint Louis, Missouri, USA. Inclusion criteria required that subjects were symptomatic and had a physician-ordered SARS-CoV-2 test performed in the course of their normal clinical care. Some subjects were enrolled prior to the return of the SARS-CoV-2 test result. Enrolled subjects who tested negative for SARS-CoV-2 are not included in the current manuscript. This report includes the first subjects enrolled in the study; however, study recruitment is ongoing. All samples were collected at the time of enrollment, which was during or immediately following evaluation in a medical facility. Patient-reported duration of illness and other clinically relevant medical information was collected at the time of enrollment from the subject, their legally authorized representative, or the medical record. The portions of the study relevant to each institution were reviewed and approved by the Washington University in Saint Louis Institutional Review Board (WU-350 study approval # 202003085) and the Missouri Baptist Medical Center Institutional Review Board (Approval # 1132). The study complied with the ethical standards of the Helsinki Declaration.

We also report findings from healthy control subjects and influenza-infected subjects enrolled in separate, ongoing studies. Control subjects had not experienced symptoms of a viral respiratory illness at the time of sample collection or within the previous 90 days, and samples were all collected before October of 2019. Influenza subjects were enrolled in the ongoing EDFLU study.(*15*) All influenza subjects were sampled in 2019 and 2020. We enrolled most influenza subjects during the course of the 2019–2020 influenza season, immediately before the spread of COVID-19 disease in the Saint Louis region. The last included influenza subject was enrolled and sampled on March 2^rd^ of 2020. The first case of COVID-19 was reported in Saint Louis on March 8^th^ of 2020 in a returning traveler. The control and influenza studies were independently approved by the Washington University Institutional Review Board (Approval #’s 201707160, 201801209, 201808171, 201710220, 201808115 and 201910011).

### Multi-Parameter Flow Cytometry

Absolute counts of CD45^+^ cells in whole blood were determined at the time of blood collection on fresh samples by flow cytometry with Precision Count Beads (BioLegend). Peripheral blood mononuclear cells (PBMCs), prepared using ficoll separation, were analyzed using a panel of antibodies directed against the following antigens: CD8 BV421 (clone RPA-T8), CD20 Pacific Blue (clone 2H7), CD16 BV570 (clone 3G8), HLA-DR BV605 (clone L243), IgD SuperBright 702 (clone IA6-2), CD19 BV750 (clone HIB19), CD45 Alexa Fluor 532 (clone HI30), CD71 PE (clone CY1G4), CD38 PE-Cy7 (clone HIT2), CD14 APC (clone M5E2), CD4 Spark 685 (clone SK3), and CD3 Alexa 700 (clone UCHT1). PBMC samples of 0.5–2×10^6^ cells were stained with a master-mix containing pre-titrated concentrations of the antibodies, along with BD Brilliant Buffer (BD Biosciences) and Zombie NIR Fixable Viability Marker (BioLegend) to differentiate live and dead cells. Samples were run on a Cytek Aurora spectral flow cytometer using SpectroFlo software (Cytek) and unmixed before final analysis was completed using FlowJo software (BD Biosciences).

### Cytokine Quantification

Plasma obtained from subjects was frozen at −80°C and subsequently analyzed using a human magnetic cytokine panel providing parallel measurement of 35 cytokines (ThermoFisher). The assay was performed according to the manufacturer’s instructions with each subject sample performed in duplicate and then analyzed on a Luminex FLEXMAP 3D instrument.

### Single-Cell RNAseq

PBMCs were suspended at 1000 cells/µL and approximately 17,400 cells were input to a 10x Genomics Chromium instrument. Each sample was used for two independent reactions, with all first reactions processed on one chip and second reactions processed on a second chip. Single-cell gene expression libraries were prepared using 5-prime (V2) kits and sequenced on the Illumina NovaSeq 6000 platform at 151×151bp. Individual libraries were processed using CellRanger (v3.1.0; 10xGenomics) with the accompanying human reference (GRCh38-3.0.0), which was modified to include the influenza A, influenza B, and COVID-19 (NC_045512.2) genomes. Processed libraries were subsequently aggregated using CellRanger, randomly subsampling mapped reads to equalize sequencing depth across cells. Filtered aggregation matrices were subsequently analyzed using Seurat(*40*) (v3.1.4), excluding cells from downstream analyses that exhibited extremes in the total number of transcripts expressed, the total number of genes expressed, or mitochondrial gene expression. For each cell we inferred cell cycle phase using markers from Tirosh et al 2016(*41*) and incorporated module scores from a number of external gene sets in the same manner.

After filtering, we first sought to characterize putative cell subsets shared across conditions by detecting integration anchors among the samples, effectively minimizing condition-associated differences. The top 2,000 variable genes were identified for each library using the “vst” method, and integration anchors were obtained using canonical correlation analysis (CCA). Data were integrated using 50 CCA dimensions and scaled to regress out the effects of total transcript count, percent of mitochondrial gene expression, and module scores associated with cell phase. Principal components (PC) were calculated and assessed for statistical significance using random permutation. The first 51 PCs (p < 0.01) were used to identify transcriptional clusters and for tSNE and UMAP dimensionality reduction. After identifying clusters on the basis of transcriptional similarities across cells from all three conditions (i.e., the “integrated” analysis), we performed pairwise differential gene expression analysis between conditions using Wilcoxon Rank Sum tests as implemented in Seurat, with default parameters. We also generated an additional UMAP projection using the top 2,000 variable genes across the entire dataset (excluding TCR and IG genes, which are known to map poorly) irrespective of the CCA but again using significant PCs; this allowed us visualize cells in a manner that did not obscure transcriptional differences owed to sample or condition but with previously identified cell subsets and transcriptional clusters from the integration analysis overlaid. We also looked within identified subsets and clusters for explicit differences in gene pathway enrichment between cells from COVID-19-infected and influenza-infected patients, COVID-19-infected and healthy patients, and influenza-infected and healthy patients. For these analyses, gene expression differences between conditions were ranked for individual subsets and transcriptional clusters by calculating differential expression under a generalized linear hurdle model(*42*). To generate gene ranks, gene-specific average log fold changes were multiplied by the absolute difference in the proportions of cells expressing the gene (+1e-4 as a lower boundary) and the inverse of the FDR-adjusted p-values, which were re-scaled from 1e-7 to 1 to institute reasonable bounds in the ranking. These ranks were used as inputs for gene set enrichment analysis(*43*) using GSEAPreranked with a classic enrichment statistic and chip-based gene collapsing based on the Human_Symbol_with_Remapping_MSigDB.v.7.0 chip(*44*). Gene sets were considered significantly enriched if they resulted in a nominal p-value < 0.05 and a q-value < 0.20.

### Data Availability

Raw flow cytometry and cytokine data can be found in Supplementary Tables 1 and 2, respectively. Single cell gene expression sequences have been uploaded to the NCBI Short Read Archive under BioProject ID PRJNA630932.

### Analysis

#### Flow Cytometry

Lymphocyte subsets were compared across healthy, influenza, and SARS-CoV-2 subjects using Mann-Whitney U tests. In the HLA-DR expression analysis, there were four negative mean fluorescence intensity observations, these were replaced with a value of 1. Results were adjusted for multiple comparisons using the Benjamin-Hochberg approach and considered significant if the adjusted p-value was less than 0.05 (FDR < 0.05).

#### Cytokines

Cytokine levels were natural safe-log transformed prior to analysis and compared across healthy, influenza, and SARS-CoV-2 subjects using Kruskal-Wallis tests. Results were adjusted for multiple comparisons using the Benjamini–Hochberg approach. Kruskall-Wallis tests with an FDR-adjusted p-value < 0.10 were selected for further investigation, and were followed up with post hoc Dunn’s tests to determine which groups differ from each other. P-values obtained from Dunn’s tests were subjected to multiple testing adjustment and considered statistically significant a FDR < 0.05.

Cytokine-cytokine co-correlations were investigated using CytoMod(*17*) utilizing both absolute and mean-adjusted (i.e., relative) cytokine values of either all COVID-19 and influenza data, using only using COVID-19 data, or only using data from hospitalized subjects of either cohort. For these correlations, values below the lower limit of detection were set to the lower limit of detection, and values above the upper limit of detection were excluded. In each instance we tested up to k = 12 modules and used gap statistic to identify the optimal k. Logistic regressions were carried out within the CytoMod framework and controlled for effects of age and the number of days after symptom onset that a sample was collected. P-values were adjusted for multiple testing as above, and all significant results reported from logistic models were assessed an FDR < 0.05.

## Supplementary Materials

Fig. S1. Gating strategy for flow cytometry analysis.

Fig. S2. Overview of cytokine levels across all subjects.

Fig. S3. Box plots of cytokine concentrations for all 35 cytokines measured.

Fig. S4. Correlations of absolute cytokine values.

Fig. S5. Modular analysis of cytokines using only samples from COVID-19 patients.

Fig. S6. Percentage of cells from each single-cell RNAseq bioinformatically-defined transcriptional clusters in COVID-19, healthy control and influenza subjects.

Fig. S7. Graphs for all results of pre-ranked gene set enrichment analysis of hallmark gene sets comparing COVID-19 and influenza subjects.

Fig. S8. Graphs for selected results from pre-ranked gene set enrichment analysis of gene ontology gene sets comparing COVID-19 and influenza subjects.

Fig. S9. Percentage of cells from each single-cell RNAseq transcriptionally identified cell subset in COVID-19, healthy control and influenza subjects.

Fig. S10. Evaluation of pre-ranked gene set enrichment analysis of gene ontology gene sets with “cortico,” “cortisol,” or “stress” in the gene set name.

S. Table 1. Raw flow cytometry data.

S. Table 2. Raw cytokine data.

## Acknowledgments

The authors would like to thank Jamie Rolando, Aaron Day, Bronson Flint, Eric Raines and support staff in the Washington University Emergency Care and Research Core for their hard work and personal dedication to collecting COVID-19 patient samples. We thank the Genome Technology Access Center (GTAC@MGI) in the Department of Genetics and McDonnell Genome Institute at Washington University School of Medicine. GTAC@MGI is partially supported by NCI Cancer Center Support Grant #P30 CA91842 to the Siteman Cancer Center and by ICTS/CTSA Grant# UL1 TR000448 from NCATS. We also thank the Bursky Center for Human Immunology and Immunotherapy Programs at Washington University, Immunomonitoring Laboratory.

## Funding

This work was supported by a grant from the Barnes Jewish Hospital Foundation to PAM, by grants NIAID R21 AI139813, U01 AI141990, Collaborative Influenza Vaccine Innovation Centers contract 75N93019C00051, and NIAID Centers of Excellence for Influenza Research and Surveillance (CEIRS) contract HHSN272201400008C to AHE and by the St. Jude Center of Excellence for Influenza Research and Surveillance (HHSN272201400006C), R01AI121832, and ALSAC to PGT. The COVID-19 clinical cohort reported in this publication was supported by the Washington University Institute of Clinical and Translational Sciences grant UL1TR002345 from the National Center for Advancing Translational Sciences (NCATS) of the National Institutes of Health (NIH). The content is solely the responsibility of the authors and does not necessarily represent the official view of the NIH.

## Author contributions

P.A.M., S.L.H., K.E.R., R.S.H., R.M.P, J.A.O., W.G.P. and A.H.E. designed and coordinated clinical studies. J.P.B., N.J.A., D.A.S., R.S.M. referred and enrolled subjects. P.A.M., J.C.C., J.S.T., A.C.M.B., P.G.T. and A.H.E. conceived of and designed the experiments performed. P.A.M., J.S.T., D.R., and D.B. performed the experiments. P.A.M., J.C.C., J.S.T., A.S., D.R., D.B., P.G.T. and A.H.E. compiled and analyzed the data. P.A.M., J.C.C., A.S., P.G.T. and A.H.E. wrote the manuscript.

## Competing interests

A.H.E. is a consultant for Inbios and Fimbrion Therapeutics. The Ellebedy laboratory received funding under sponsored research agreements from Emergent BioSolutions. All other authors declare that they do not have any commercial interests or other associations that pose a conflict of interest with the published work.

## Data and materials availability

Single cell gene expression sequences have been uploaded to the NCBI Short Read Archive under BioProject ID PRJNA630932. Requests for data or materials to: Philip A. Mudd and Paul G. Thomas and Ali H. Ellebedy.

## Supplementary Materials

**Supplementary Figure 1**. Gating strategy for flow cytometry analysis of myeloid **(A)** and lymphoid **(B)** subsets. **(C)** Expression of HLA-DR, measured by mean fluorescence intensity, on B cells, CD8 T cells, and CD4 T cells.

**Supplementary Figure 2**. Overview of cytokine levels across all subjects, data points from the 3 COVID-19 patients with extremely high cytokine concentrations are shown in red.

**Supplementary Figure 3**. Box plots of cytokine concentrations in COVID-19, influenza, and healthy subjects for each of the 35 cytokines measured. P-values presented are raw, unadjusted Kruskal-Wallis tests or Mann-Whitney U tests.

**Supplementary Figure 4**. Correlations of absolute cytokine values are visualized after hierarchical clustering (left), as well as the proportion of times cytokine pairs clustered together during hierarchical clustering over 1,000 permutations (right) for: **(A)** all samples from COVID-19- and influenza-infected patients, **(B)** only samples obtained from COVID-19-infected patients, and **(C)** only samples obtained from hospitalized patients. Data from healthy patients were not included in these analyses. Figures were produced using CytoMod.

**Supplementary Figure 5**. Modular analysis of cytokines as described in the manuscript were repeated using only samples from COVID-19-infected patients. **(A)** Odds ratio heatmap from logistic regression analysis testing for associations between cytokines/cytokine modules and disease severity (as defined by presentation that resulted in intubation or death, “IntOrDec”). Only statistically significant odds ratios are visualized; all non-grey values correspond to statistically significant results after multiple testing adjusted to ensure a false discovery rate <0.05. Asterisks indicate an additional, more stringent Bonferroni multiple testing adjustment, with *** corresponding to FWER (family-wise error rate) < 0.001 and ** corresponding to FWER < 0.01. Odds ratios are presented in a color scales that spans from dark purple (OR = 1/2.5), corresponding to a negative association between cytokine expression and severity, to dark brown (OR = 2.5), corresponding to a positive association between cytokine expression and severity. Modules were defined by hierarchical clustering of cytokine-cytokine correlations **(B)**. **(C)** visualizes the proportion of times cytokine pairs clustered together during hierarchical clustering over 1,000 permutations. Figures were produced using CytoMod.

**Supplementary Figure 6**. For each bioinformatically defined transcriptional cluster, the percentage of cells from each single-cell gene expression library that were assigned to that cluster is presented. Colors correspond to the patient from which the sample was obtained, with each patient represented by two independent gene expression libraries constructed from distinct cell aliquots from the same PBMC sample.

**Supplementary Figure 7**. Graphs depicting all results of Preranked Gene Set Enrichment Analysis of Hallmark gene sets when comparing COVID-19-infected and influenza-infected subjects on a per-cell-subset basis. Red: significantly enriched in cells from COVID-19-infected patients; blue: significantly enriched in cells from influenza-infected patients; grey: not statistically enriched in either condition.

**Supplementary Figure 8**. Graphs depicting selected results from Preranked Gene Set Enrichment Analysis of Gene Ontology gene sets. Gene Ontology Biological Processes gene sets that passed default GSEA thresholds were tested, with results adjusted for multiple testing, and all results with the term “interferon” in the gene set name that were significant in either direction for at least one cell subset are plotted. Red: significantly enriched in cells from COVID-19-infected patients; blue: significantly enriched in cells from influenza-infected patients; grey: not statistically enriched in either condition.

**Supplementary Figure 9**. For each transcriptionally identified cell subset, the percentage of cells from each single-cell gene expression library that were assigned to that subset is presented. Colors correspond to patient from which the sample was obtained, with each patient represented by two independent gene expression libraries constructed from distinct cell aliquots from the same PBMC sample.

**Supplementary Figure 10**. Graphs depicting selected results from Preranked Gene Set Enrichment Analysis of Gene Ontology gene sets. Gene Ontology Biological Processes gene sets that passed default GSEA thresholds were tested, with results adjusted for multiple testing, and all results with the terms “cortico”, “cortisol”, or “stress” in the gene set name that were significant in either direction for at least one cell subset are plotted. Red: significantly enriched in cells from COVID-19-infected patients; blue: significantly enriched in cells from influenza-infected patients; grey: not statistically enriched in either condition.

Additional supplemental data files Supplementary Table 1 and Supplementary Table 2 are available in the online version.

## Notes

### Author Declarations

Washington University in Saint Louis School of Medicine Institutional Review Board Approval #s 202003085, 201707160, 201801209, 201808171, 201710220, 201808115 and 201910011. Missouri Baptist Medical Center Institutional Review Board Approval #1132.

